# A longitudinal investigation of blood neurofilament light chain levels in chronic cocaine users

**DOI:** 10.1101/2022.02.03.22270384

**Authors:** F. Bavato, A.K. Kexel, B. Kluwe-Schiavon, A. Maceski, M.R Baumgartner, E. Seifritz, J. Kuhle, B.B. Quednow

**Author notes:** **Corresponding author:** Francesco Bavato, MD, Department of Psychiatry, Psychotherapy and Psychosomatics, Psychiatric Hospital, University of Zurich, Lenggstrasse 31, CH-8032 Zurich, Switzerland. **Declaration of interest:** None. **Author agreement:** All authors agreed with the current version of the manuscript and approved the submission to the Journal.

## Abstract

**Objectives:** To explore the hypothesis that plasma levels of neurofilament light chain (NfL), a marker of neuroaxonal pathology, are elevated in chronic cocaine users (CU) and longitudinally associated with changes in cocaine use.

**Methods:** As part of the Social Stress Cocaine Study (SSCP), we assessed 35 CU and 35 stimulant-naïve healthy controls (HC) at baseline and at a 4-month follow-up. Plasma NfL levels were determined from blood samples using single molecule array (SIMOA) technology. Substance use was subjectively assessed with an extensive interview and objectively measured via toxicological analysis of urine and 4-month hair samples.

**Results:** In a generalized linear model corrected for sex, age, and body mass index, NfL plasma levels were elevated in CU compared to HC (p<0.05). A moderate positive correlation between cocaine hair concentration and NfL levels was also found in CU (r(s)=0.36, p=0.03). Changes in cocaine hair concentration (group analysis of increasers vs. decreasers) over the 4-month interval predicted NfL levels at follow-up (p=0.002), indicating a rise in NfL with increased cocaine use and a reduction with decreased use. No associations between use or change of use of other substances (including the cocaine adulterant levamisole) and NfL levels were found (r(s)≤±0.27, p>0.05).

**Conclusions:** Our findings demonstrate that NfL is a sensitive marker for assessing cocaine-related brain pathology, supporting the utility of blood NfL analysis in addiction research. The results also suggest that cocaine use should be considered a potential confounder in diagnostic applications and clinical studies using NfL.

## Introduction

Chronic cocaine use has been consistently associated with a range of structural brain alterations, including volumetric changes in cortical and subcortical regions and disruption of white matter tracts.^1-5^ Although some recent longitudinal studies have suggested that neuroanatomical alterations in fronto-cortical areas covary with changes of cocaine consumption,^6,7^ most of the findings are limited to cross-sectional observations. Thus, the extent to which these structural impairments are directly related to cocaine-induced brain pathology (neurotoxicity) rather than preexisting traits (predisposition) remains unclear.^8,9^ Moreover, cocaine adulteration with neurotoxic compounds (e.g., the anthelmintic levamisole),^10,11^ concomitant use of other illicit substances or alcohol,^12^ and comorbidity with psychiatric disorders (e.g., depression)^12-15^ might influence brain integrity in chronic cocaine users (CU) independently of cocaine intake itself. Considering the limitations of current diagnostic methods, which are mostly limited to cost-intensive neuroimaging measures, the introduction of new markers of active brain pathology may help clarify crucial questions on substance-induced brain pathology and provide new monitoring tools for physicians working in addiction medicine.^16^

Neurofilament light chain (NfL) is a cytoskeletal protein that is released in extracellular matrices during neuroaxonal damage.^17^ Since the recent introduction of highly sensitive assay methods, NfL has emerged as the most promising blood marker of neuroaxonal pathologies for clinical use.^18^ In particular, NfL levels have been shown to reflect active brain disease, dynamically respond to therapeutic interventions, and offer prognostic information in a number of neuropsychiatric disorders, such as multiple sclerosis, neurodegenerative disorders, traumatic brain injuries, and depression.^14,19,20^ Our recent work also reported elevated NfL levels in patients with chronic ketamine addiction, demonstrating its sensitivity to substance-related brain pathology.^13^

Although cocaine use has been associated with several structural brain changes, this is the first study to investigate its impact on NfL levels. We compared the NfL levels of a sample of 35 CU with 35 healthy controls (HC) at baseline. Additionally, we investigated the dose–response relationship between NfL concentrations and cocaine consumption at baseline. Finally, we evaluated longitudinal changes in NfL levels in relation to objectively confirmed changes in cocaine consumption across a 4-month follow-up period as determined by hair testing. Based on previous cross-sectional findings linking structural brain alterations with chronic cocaine use,^1-4^ we hypothesized that NfL levels would be elevated in CU at baseline in a dose-dependent manner. Moreover, we expected that NfL levels at follow-up would be associated with changes in cocaine consumption during the 4-month interval, as previously shown for anatomical and cognitive measures in cocaine user populations.^7,21,22^ Finally, we expected that levamisole (a common adulterant of cocaine) would have an additional impact on NfL levels, given its proven neurotoxic effects on white and gray matter.^10,11,23,24^

The identification of a blood marker of active brain pathology that is sensitive to substance-induced neurotoxicity and dynamically responds to longitudinal changes in substance intake would substantially improve clinical monitoring in the field of substance use and addiction. Moreover, considering the increasing attention NfL analysis is receiving in neurology and psychiatry, the identification of variables that confound NfL levels is crucial for its use as a marker in clinical settings.

## Methods

### Standard protocol approvals, registrations, and patient consent

This study was approved by the Research Ethics Committee of the Canton of Zurich (BASEC ID 2016-00278 and 2021-01853). All participants provided written informed consent prior to their enrollment in the study and were financially compensated for their participation. Clinical and laboratory investigations were conducted in strict accordance with the principles of the Declaration of Helsinki.

### Participants

The data were collected in the context of the Social Stress Cocaine Study (SSCP) at the Psychiatric Hospital of the University of Zurich.^15,25^ The general exclusion criteria were a family history of genetically transmitted psychiatric disorders (h^2^>0.5, e.g., autism, schizophrenia, or bipolar disorder); any severe neurological disorder or brain injury; a current diagnosis of infectious disease or severe somatic disorder; a history of autoimmune, endocrine, or rheumatoid arthritis; intake of medication with potential action on the central nervous system or the physiological stress system during the previous three days; participation in a large previous study conducted by our lab, the Zurich Cocaine Cognition Study;^7,10^ and (for women) being pregnant or breastfeeding. The criteria for inclusion of CU in the study were cocaine as the primary substance of use; a lifetime cumulative consumption of at least 100 g of cocaine, estimated by self-report; and a current abstinence duration of <6 months. We also excluded CU who regularly used illegal substances other than cocaine, such as heroin or other opioids (with the exception of irregular cannabis use); those with a polysubstance use pattern according to DSM-IV-TR; and those with a DSM-IV-TR axis I adult psychiatric disorder diagnosis other than cocaine, cannabis, or alcohol abuse or dependence, previous depressive episodes, and attention deficit hyperactivity disorder (ADHD). Stimulant-naïve HC were matched for sex. The exclusion criteria for HC were a DSM-IV-TR axis I adult psychiatric disorder or recurrent illegal substance use (>15 occasions in the lifetime, with the exception of irregular cannabis use).

For the current investigation, a sample of 37 CU and 39 HC was initially available. However, in accordance with our exclusion criteria, two CU were retrospectively excluded because of polytoxic substance use patterns and low cocaine consumption, confirmed by toxicological hair analysis. Four HC were also excluded because of positive hair toxicology for cocaine or other substances according to published thresholds,^26^ which is in line with recent prevalence rates among young adults in the Zurich area.^27^ Therefore, a sample of 35 CU and 35 HC was considered for the final analyses.

### Assessments

#### Procedure

The test procedure was similar at baseline (T1) and follow-up (T2) visits. The interval between T1 and T2 was approximately 4 months (number of days between T1 and T2, mean±SD: HC=135±19; CU=169±71). Both visits included a clinical and substance-related assessment, blood sampling for NfL analysis, and toxicological analysis of urine and hair.

#### Clinical and substance use assessment

The psychopathological assessment was carried out by trained psychologists using the Structured Clinical Interview I (SCID-I) in accordance with DSM-IV-TR.^28^ Depressive symptoms were assessed using the Beck Depression Inventory (BDI).^29^ The Childhood Trauma Questionnaire (CTQ) was used to screen for traumatic childhood experiences.^30^ Symptoms of ADHD were assessed using the ADHD self-rating scale (ADHD-SR).^31^ Self-reported substance use was assessed with the structured and standardized Interview for Psychotropic Drug Consumption (IPDC).^32^

#### Neurofilament light chain analysis

Plasma NfL levels were measured using single-molecule array (SIMOA) technology at the University Hospital Basel. Blood was collected in EDTA tubes and directly centrifuged for 15 min at 1500 g at room temperature. All plasma samples were frozen and stored at −80°C. All intra-assay coefficients of variation of duplicate determinations obtained by SIMOA analysis were below 15%. The mean coefficient of variation was 4.78% in the CU group and 6.25% in the HC group. To minimize the possibility of technical issues or batch effects, T1 and T2 samples were analyzed together at the end of the study.

#### Urine and hair toxicological analysis

Urine analyses using a semi-quantitative enzyme multiplied immunoassay method targeted the following substances: amphetamines, barbiturates, benzodiazepines, cocaine, methadone, morphine-related opiates, and delta-9-tetrahydrocannabinol. In addition, quantitative analysis of hair samples using liquid chromatography tandem mass spectrometry (LC-MS/MS) was conducted to investigate substance consumption over the previous 4 months, as represented in the proximal 4 cm-segment of the hair samples taken from the occiput. In total, 88 substances and substance metabolites were assessed.^33^ The parameter cocaine_total_ (= Cocaine + Benzoylecgonine + Norcocaine) was calculated. Together with the corresponding metabolic ratios, cocaine_total_ offers a robust procedure for discriminating between incorporation and contamination of hair.^34^

### Statistical analysis

#### Preliminary analysis

All statistical analyses were computed with SPSS version 25 (IBM Corp., SPSS Inc., Chicago, IL, USA) and R-Studio (Version 3.6.1). Quantitative variables were tested for normal distribution using the Kolmogorov–Smirnov test. Between-group comparisons (CU/HC) of sociodemographic and clinical data and substance use variables were performed using Student’s t-tests (for normally distributed quantitative data), Mann–Whitney U-tests (for non-normally distributed quantitative data), or Pearson’s chi-square test (for categorical data). The significance level was set at p<0.05 (two-tailed).

#### Effects of cocaine intake on NfL levels at baseline

As a first step, we performed a generalized linear model (GLM) with Gaussian distribution and log link function to test group effects on NfL levels. We included group (CU/HC) and sex (female/male) as fixed factors and age and BMI as further covariates. The model selection was based on lognormal distribution of NfL levels. Age was included due to the strong link between advancing age and increased NfL levels,^35^ and BMI was included due to its potential confounding effects on NfL levels.^35^ Sex was included to correct for potential sex-specific effects of cocaine use on brain integrity^3^. As a second step, Spearman’s correlation coefficients were calculated to investigate the dose–response relationships between substance use and NfL levels. Hair concentrations of substances and self-reported substance use variables were used for the correlation analysis.

#### Longitudinal effects of changes in cocaine use on NfL levels at follow-up

At follow-up, CU were characterized as either increasers (participants who elevated their cocaine use between T1 and T2) or decreasers (participants who reduced their cocaine use in the same period). The assignment criteria were based on absolute changes in cocaine concentration in toxicological hair analysis between T1 and T2. The effect of changes in cocaine consumption on NfL levels at follow-up was analyzed using a GLM with Gaussian distribution and log link function. We included group (increasers-/decreasers) and sex (female/male) as fixed factors and age, BMI, and NfL levels at baseline as further covariates. As a second step, Spearman’s correlation coefficients were calculated to investigate dose–response effects of changes in substance use on changes in NfL levels. For this analysis, Δ-values were calculated to define changes in objective variables of illicit substance use and self-reported alcohol intake (i.e., Δ_COCAINE_=[cocaine concentration in hair at T2]–[cocaine concentration in hair at T1]) and NfL (Δ_NfL_=[NfL level at T2]–[NfL level at T1]). Extreme outliers were detected using the 3xIQR detection rule on Δ_NfL_ values and excluded from the longitudinal analysis. We applied the 3xIQR detection rule, as it is a conservative and robust method for small samples with symmetric distributions.^36^ Data from participants, who did not take part to the T2 visit, were excluded from longitudinal analysis.

### Data availability

Anonymized data will be shared by request with any qualified investigator with an institutional review board approval for the purposes of validation and/or replication using our center’s established procedures for sharing data.

## Results

### Sociodemographic and clinical data and substance use patterns

The sociodemographic and clinical data of the 35 CU and 35 HC are summarized in Table 1. Age and BMI score were both higher in CU compared to HC and were therefore included as cofactors in the GLM analysis investigating group effect on NfL values. We found that CU had fewer years of school than HC, which is in line with findings in other cocaine user populations.^37^ As expected, CU scored significantly higher in the BDI and ADHD-SR and had a higher frequency of major depressive disorder (MDD) diagnosis than HC (according to DSM criteria).^37^ Substance use patterns are summarized in the supplementary materials (Table S1).

**Table 1.** Sociodemographic, clinical characteristics and NFL levels at baseline.

| Variables | Cocaine users (N=35) | Healthy controls (N=35) | Test statistics | df | *P value* |
| --- | --- | --- | --- | --- | --- |
| Age, years | 33.3 ± 7.3 | 29.49 ± 7.1 | U=397.5 | - | **0.011** |
| Sex (f/m) | 11/24 | 13/22 | x<sup>2</sup>=0.25 | 1 | 0.802 |
| BMI, kg/m<sup>2</sup> | 25.2 ± 4.0 | 23.3 ± 3.2 | t=2.20 | 68 | **0.032** |
| School education, years | 9.6 ± 1.1 | 10.5 ± 1.5 | U=450.5 | - | **0.031** |
| Family psychiatric history (yes/no) | 3/32 | 0/35 | x<sup>2</sup>=3.13 | 1 | 0.239 |
| ADHD_SR, score | 15.0 ± 10.5 | 9.6 ± 7.8 | U=417.5 | - | **0.022** |
| ADHD DSM (yes/no)<sup>a</sup> | 9/26 | 5/30 | x<sup>2</sup>=1.45 | 1 | 0.371 |
| History of MDD DSM (yes/no)<sup>b</sup> | 14/21 | 3/32 | x<sup>2</sup>=9.40 | 1 | **0.004** |
| CTQ, score | 53.0 ± 14.8 | 47.1 ± 9.4 | U=484.5 | - | 0.132 |
| BDI, score | 7.9 ± 6.8 | 4.1 ± 5.4 | U=365.0 | - | **0.003** |

Table reports counts or means ± standard deviations. Significant group differences are shown in bold. t = Student t-test; x<sup>2</sup> = Pearson chi-square; U = Mann-Whitney test.
<sup>a</sup> Cut-off according to DSM-IV criteria as assessed by the ADHD-SR questionnaire. <sup>b</sup> Cut-off according to DSM-IV criteria as assessed by SCID-I interview.
Abbreviations: ADHD, Attention deficit hyperactivity disorder; BDI: Beck Depression Inventory; BMI: Body Mass Index; CTQ: Childhood Trauma Questionnaire; MDD: major depressive disorder.

In accordance with the inclusion criteria, hair samples confirmed a clear dominance of cocaine use over all other illegal substances. The mean cocaine concentration in hair (mean±SD: cocaine_total_=33,102±67,934 pg/mg) was also in line with previous findings in dependent CU obtained by our group.^37^ In contrast, the mean concentration of levamisole (a neurotoxic cocaine adulterant) in hair (mean±SD: 2,962±9,604 pg/mg) and the mean levamisole–cocaine ratio in hair (0.17) were substantially lower than in our previous reports.^10^ However, this finding is coherent with the drop in levamisole prevalence observed in Switzerland during the period of recruitment (2017–2018).^10^

### Effects of cocaine intake on NfL levels at baseline

Unadjusted mean NfL levels in plasma were significantly higher in CU than in HC (mean/median [interquartile range (IQR)] in pg/ml: 6.75/5.70 [4.00–6.40] vs. 4.85/4.40 [3.05–5.75]; U=414.00, p=0.020), with medium effect size (Cohen’s d=0.51) (see Figure 1). In the correlation analysis, a positive association of age with NfL levels (r(s)=0.510, p=0.002) and a negative association of BMI with NfL levels (r(s)=−0.430; p=0.010) were observed in HC, which is consistent with previous findings^35^ (supplementary Table S2). In contrast, we found no significant correlation of any sociodemographic or clinical variable with NfL levels in CU (r(s)≤±0.33, p>0.05). In the GLM analysis accounting for age, sex, and BMI, a significant group effect confirmed higher NfL plasma levels in CU at baseline (see Table 2). Age was a significant covariate, while BMI showed a trend level negative association with NfL. Sex had no significant impact on the model. To further exclude the potential impact of demographic variables on elevated NfL levels in CU, we also compared NfL levels in 22 CU and 22 HC (from the same sample) who were matchedfor age (±2years) and did not differ in mean BMI.. Again, we found significantly higher NfL levels in CU than in HC (Z=−2.22, p=0.026). At baseline, we found no association between NfL levels and subclinical symptoms of depression, ADHD, or childhood trauma (see Table S2). However, NfL was positively associated with hair concentration of cocaine_total_ (r(s)=0.364, p=0.032) (see Figure 1). No significant association was found between NfL and levamisole, other illicit substances, or weekly alcohol intake (see Table S3) (r(s)≤±0.27, p>0.05). Moreover, NfL was not associated with recent cocaine use assessed by urine toxicology (r(s)=0.02, p=0.91).

**Table 2.**
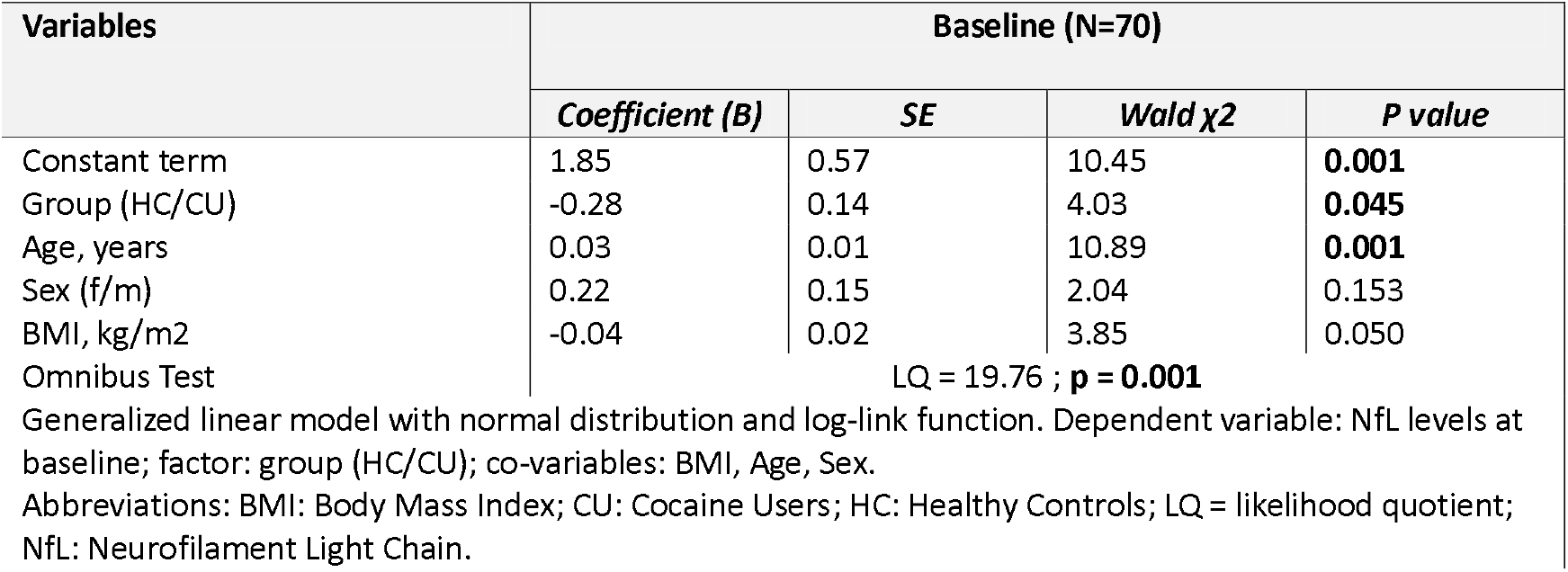
Generalized linear model for sociodemographic and clinical characteristics predicting NfL levels at baseline.

| Variables | Baseline (N=70) |  |  |  |
| --- | --- | --- | --- | --- |
| | Coefficient (B) | SE | Wald $\chi^2$ | P value |
| Constant term | 1.85 | 0.57 | 10.45 | **0.001** |
| Group (HC/CU) | -0.28 | 0.14 | 4.03 | **0.045** |
| Age, years | 0.03 | 0.01 | 10.89 | **0.001** |
| Sex (f/m) | 0.22 | 0.15 | 2.04 | 0.153 |
| BMI, kg/m2 | -0.04 | 0.02 | 3.85 | 0.050 |
| Omnibus Test | LQ = 19.76 ; **p = 0.001** |  |  |  |
| Generalized linear model with normal distribution and log-link function. Dependent variable: NfL levels at baseline; factor: group (HC/CU); co-variables: BMI, Age, Sex. |  |  |  |  |
| Abbreviations: BMI: Body Mass Index; CU: Cocaine Users; HC: Healthy Controls; LQ = likelihood quotient; NfL: Neurofilament Light Chain. |  |  |  |  |

**Figure 1.**
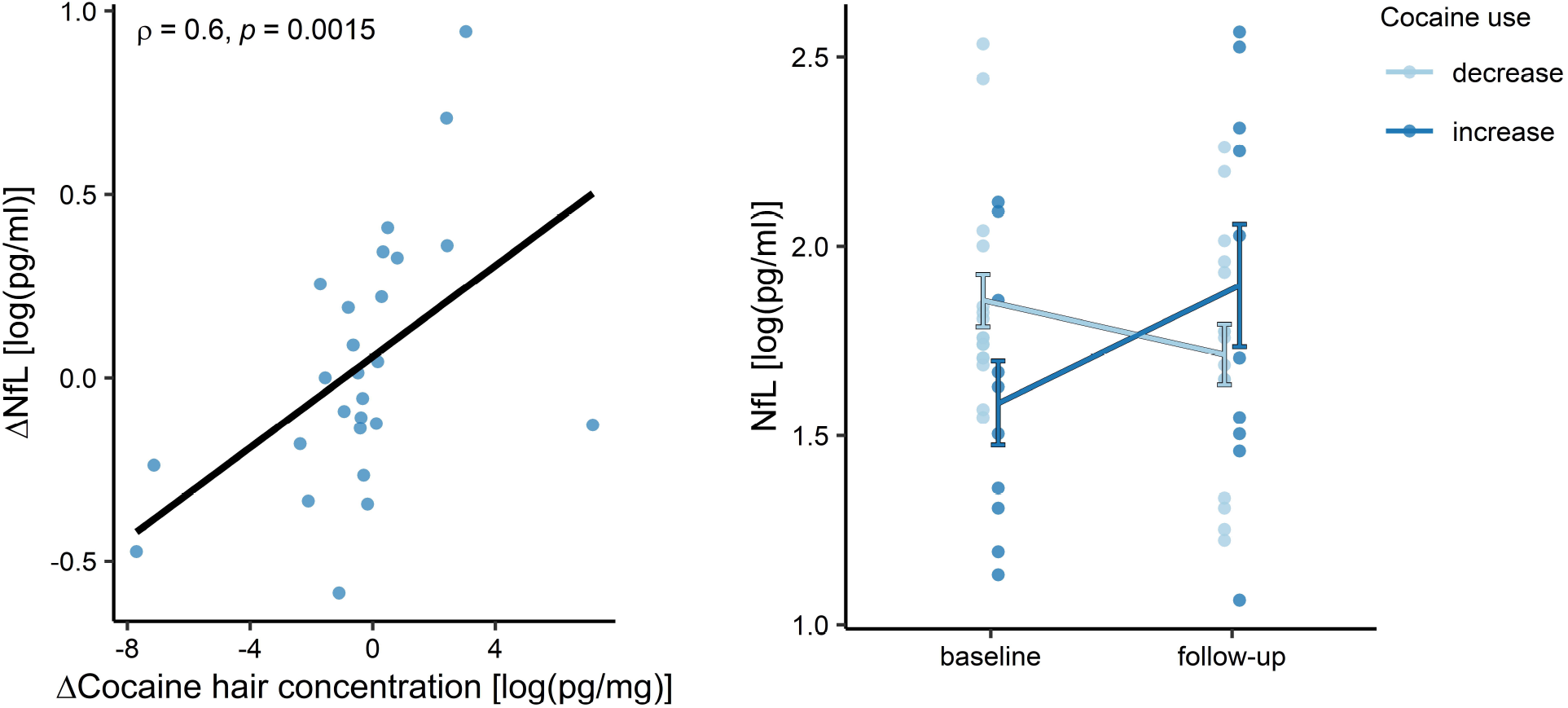
NfL levels in healthy controls and chronic cocaine users. *Left panel*: Boxplots showing individual plasma NfL levels (log-transformed). Central horizontal lines indicate median values; boxes illustrate the ranges between lower and upper quartiles. *Right panel:* Scatterplot showing the relationship between hair concentration of cocaine_total_ and plasma NfL levels at baseline (both log-transformed). Cocaine_total_ (= Cocaine + Benzoylecgonine + Norcocaine) is, together with the corresponding metabolic ratios, a more robust procedure for discrimination between incorporation and contamination of hairs. *Abbreviations:* HC: Healthy Controls; CU: Cocaine Users; NfL: Neurofilament Light Chain.

### Longitudinal effects of increased/decreased cocaine consumption on NfL levels at follow-up

At follow-up, 30 of 35 CU and 29 of 35 HC were available to be re-tested. The dropouts resulted from participants, who did not respond to the invitation for the T2 visit. Unadjusted mean NfL levels at follow-up were significantly higher in CU than in HC (mean/median [IQR] in pg/ml: 7.75/5.85 [3.42– 8.28] vs. 5.03/4.90 [3.33–6.48]; U=280.50; p=0.019), with a large effect size (Cohen’s d=0.75) (see Figure 1). In the GLM analysis of follow-up values, we observed a significant effect of group (CU/HC) on NfL at follow-up (B=−0.39, SE=4.73, p=0.030).

Regarding cocaine levels in hair, cocaine increasers (n=11) showed a mean increase of cocaine_total_ concentration of +10,296±13,633 pg/mg (mean change, percent: +441%; absolute change range: +475 pg/mg to +37,125 pg/mg), whereas cocaine decreasers (n=19) showed a mean decrease of cocaine_total_ of −31,280±60,877 pg/mg (mean change, percent: −56%; absolute change range: −570 pg/mg to −232,200 pg/mg). Four subjects were excluded from the longitudinal analysis, as they showed extreme variations in NfL values (above 3IQR) between T1 and T2. In the GLM adjusted for sex, age, BMI, and NfL levels at baseline, we found a significant group effect of change in cocaine use on NfL levels at follow-up (p<0.001) (see Figure 2). In particular, Δ_NfL_ ([NfL level at T2]–[NfL level at T1]) was significantly elevated in increasers (mean±SD: 2.29±2.50 pg/ml; T=2.90, p=0.018) and reduced in decreasers (mean±SD: −0.85±1.41 pg/ml; T=−2.407, p=0.029) but showed no significant change in HC (mean±SD: 0.09±1.25 pg/ml; T=0.385, p=0.703). In addition, NfL levels at baseline were positively associated with NfL levels at follow-up, while age, sex, and BMI had no significant impact on the model (see Table 3). Longitudinal dose–response effects of cocaine intake on NfL levels were demonstrated by a positive association of Δ_COCAINE_ in hair with Δ_NfL_ (r(s)=0.58 p=0.002) (Figure 2). In contrast, neither changes in hair concentration of other illegal substances or levamisole nor changes in alcohol were associated with changes in NfL values (see Table S4).

**Table 3.** Generalized linear model for sociodemographic and clinical characteristics predicting NFL levels at 4-months-follow-up in cocaine users.

| Variables | Cocaine users (N= 26) |  |  |  |
| --- | --- | --- | --- | --- |
| | Coefficient (B) | SE | Wald $\chi^2$ | P value |
| Constant term | 1.63 | 0.59 | 7.67 | **0.006** |
| Group (increasers/decreasers) | -0.48 | 0.12 | 15.14 | **>0.001** |
| NFL levels at baseline, pg/ml | 0.12 | 0.03 | 18.75 | **>0.001** |
| Age, years | 0.00 | 0.01 | 0.16 | 0.686 |
| Sex (f/m) | -0.15 | 0.17 | 0.82 | 0.366 |
| BMI, kg/m2 | -0.01 | 0.02 | 0.37 | 0.541 |
| Omnibus Test | LQ =21.80; **p = 0.001** |  |  |  |
| Generalized linear model with normal distribution and log-link function. Dependent variable: NfL levels at follow-up; factor: group (increasers/decreasers); co-variables: NfL levels at baseline, BMI, Age, Sex. |  |  |  |  |
| Abbreviations: BMI: Body Mass Index; LQ = likelihood quotient; NfL: Neurofilament Light Chain. |  |  |  |  |

**Figure 2.**
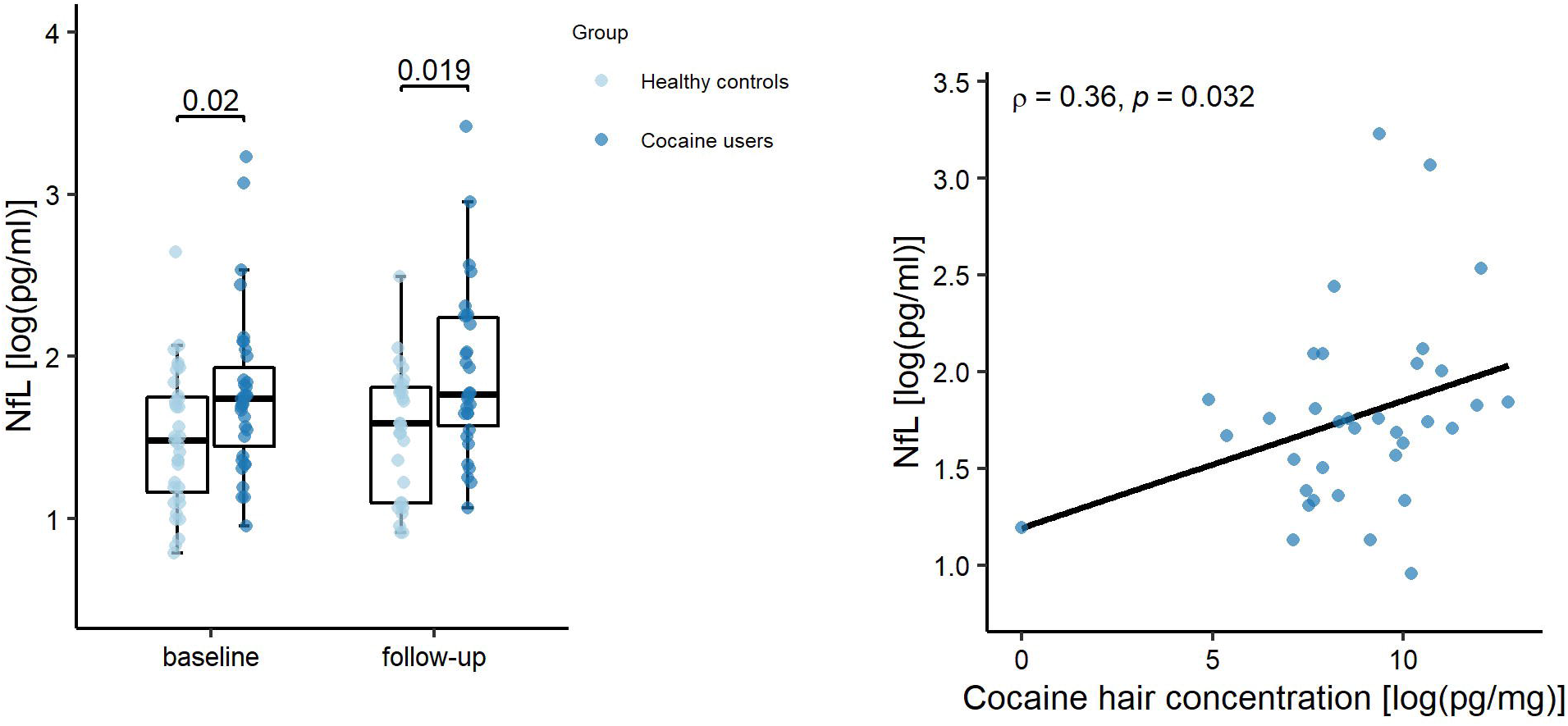
Longitudinal changes of NfL levels depending on variation of cocaine use. *Left panel:* Scatterplot showing the relationship between variations of hair concentration of cocaine_total_ and plasma NfL levels over the follow-up period. Delta values are calculated between timepoints ((ΔX=[value X at T2]-[value X at T1]). *Right panel:* Boxplots showing longitudinal variations of NfL levels in chronic cocaine users with decreased vs. increased hair concentration of cocaine over the follow-up period. Mean values ± SD of NfL levels are indicated. *Abbreviations:* NfL: Neurofilament Light Chain.

## Discussion

The aim of the present study was to investigate a potential association between chronic cocaine use and NfL levels in blood using a longitudinal design to assess cocaine-related neurotoxicity. We found that plasma NfL levels were higher in CU than HC and were positively associated with cocaine use intensity, as confirmed by hair toxicology. Moreover, NfL levels at the 4-month follow-up were predicted by objectively verified changes in cocaine intake during the interval period, showing that increased cocaine use was associated with elevated NfL levels, while reduced use was associated with lower NfL concentrations.

The findings suggest that cocaine use may impact neuroaxonal structures. In particular, NfL elevation in CU was positively associated with increased cocaine intake, as confirmed by hair analysis, which makes it an objective, reliable, and effective way of discriminating between cocaine ingestion and external contamination.^38^ In contrast, we found no association (cross-sectional or longitudinal) between NfL and intake of other illicit substances or alcohol in our sample. However, due to our preselection of primary cocaine users, we cannot rule out potential neurotoxic effects of other substances. Contrary to our expectations, NfL was not associated with the concentration of the adulterant levamisole, which has been linked to structural brain alterations in CU.^10,11,23,24^ The findings of previous studies by our group suggested that levamisole induces thinning of the prefrontal cortex and increases white matter pathology.^10,11^ However, in the current sample, the concentration of levamisole in hair was relatively low compared to our previous studies, which is in line with the observed reduction of levamisole concentration in cocaine samples in Switzerland during the recruitment period.^10^ Similarly, we found no link between NfL levels and depressive symptoms, history of childhood trauma, or ADHD symptoms. Higher NfL levels have previously been described in patients with MDD and in a sample of treatment-seeking ketamine users with a history of MDD.^13,14^ However, in this study, we specifically excluded subjects with current major depressive episodes. Thus, the lack of associations between NfL levels and depressive symptoms in the present study is consistent with the view that neuroaxonal pathology in MDD is strongly stage-dependent.^14^ Finally, NfL levels at follow-up were predicted by an interaction between baseline levels and changes in cocaine use over the interval period. This finding suggests that elevated NfL levels in CU may reflect direct substance-induced effects on neuroaxonal structures rather than a preexisting predisposition. This is consistent with evidence from patients with neuroinflammatory disorders, which demonstrated that NfL increases reflect active (stage-dependent) rather than preexisting (trait-dependent) brain pathology and that NfL levels may normalize with therapeutic interventions.^39^

The elevation of NfL levels observed in CU is in line with clinical and preclinical evidence of cocaine-induced neurotoxicity.^4,40^ However, the lack of specificity of the NfL response does not allow us to make clear assumptions regarding the specific neuropathological processes involved in its elevation. In preclinical models, repeated cocaine exposure was shown to activate several neuropathological pathways (i.e., oxidative stress reaction, mitochondrial dysfunction, excitotoxicity, and autophagy), mostly mediated by long-term neurometabolic dysregulations.^40,41^ The effects of repeated cocaine administration included alterations of cytoskeletal structures (e.g., neurofilament proteins) and were linked to neuronal adaptations of dopaminergic circuits.^42,43^ Post-mortem investigations also suggested damage to dopaminergic neurons in the basal ganglia and white matter disruption in CU.^4,44^ Neuroimaging studies have found reductions in gray matter volumes, especially in the frontal and insular areas,^1,3,5^ and widespread alterations of major white matter tracts in CU.^4^ These structural changes were linked to cognitive dysfunctions and seemed to be partially reversible after longer periods of abstinence or decreased cocaine use.^2,7^ In particular, recovery potential was observed for gray matter volume in the inferior frontal gyrus and the ventromedial prefrontal region^6^ and for cortical thickness in the frontal lobe area.^7^ Notably, previous studies have demonstrated the susceptibility of NfL levels to microstructural brain alterations that are relevant for cognitive performance have been already demonstrated in previous studies in patients with psychiatric disorders and even in healthy controls.^14,45^ Moreover, our findings are consistent with previous reports on the reversibility of neuroanatomical alterations in fronto-cortical brain areas.^7^ Thus, repeated cocaine exposure may induce microstructural adaptations in fronto-cortical regions via neurometabolic dysregulations, resulting in peripheral NfL elevation and cognitive dysfunctions. A reduction of cocaine consumption might then partially restore neurometabolic homeostasis and allow structural recovery, specifically in frontal areas, resulting in normalization of NfL values. In contrast, increased cocaine consumption would likely result in continued neurometabolic dysregulation and induce further structural damage and NfL release.

Cocaine use has also been related to cerebrovascular disease via multiple pathways, including direct (vasospasm, cerebral vasculitis) and indirect (hypertensive surges, arrhythmias, induction of plaque growth, and enhanced platelet aggregation) mechanisms.^46^ Potential acute manifestations, such as ischemic or hemorrhagic stroke, are accompanied by more subtle and chronic alterations, such as small vessels pathology with white matter hyperintensities.^46^ Thus, NfL elevation in CU might also be (at least partially) caused by subclinical cerebrovascular pathology, as observed in patients with clinically silent small vessel disease.^47^ However, the findings of a previous investigation by our group suggested that the extension of white matter hyperintensities may be associated more with cocaine adulteration with levamisole than with the level of cocaine intake.^11^ Thus, considering the longitudinal response of NfL levels to changes in cocaine intake and the lack of association between NfL and levamisole, it is reasonable to assume that NfL may reflect cocaine-induced microstructural alterations rather than cerebrovascular disease.

Overall, the identification of a peripheral marker that is (1) minimally invasive, widely available, and methodically robust and (2) sensitive enough to detect substance-induced or at least substance-related brain pathology and (3) dynamically respond to variations in cocaine intake over a relatively short period (weeks to months) could be of significant benefit for the addiction field. For instance, it could be used to estimate the severity of neuropsychiatric sequelae of substance use, which could be useful for motivating patients to enter or intensify treatment. Not only could NfL analysis improve our understanding of substance-related brain damage, but it could also have relevant applications in clinical settings. Based on our findings, NfL appears to be generally suitable for monitoring the biological harm caused by different psychoactive substances and their adulterants. It may also be useful for identifying clinical factors involved in vulnerability or resilience to substance-related brain impairment, assessing the impact of therapeutic interventions on substance users, and facilitating personalized treatments. Moreover, considering the prevalence of cocaine use and the overall use of illicit stimulants in the general population, our study warrant consideration on illicit substance use as a potential confounding factor influencing NfL investigations in cohort studies with primary neurological or psychiatric diseases.

The present study has certain limitations that must be acknowledged. First, the sample size was moderate. However, by performing detailed assessments of substance use patterns and objective quantifications of substance intake by toxicological hair analysis, strictly excluding participants with neuropsychiatric comorbidities or severe polysubstance use, and adopting a longitudinal design, we were able to provide robust evidence of cocaine-induced effects on NfL levels. Second, the lack of neuroimaging data limits our speculations on the neuroanatomical correlates of NfL elevations and should be considered in future investigations. Third, we did not systematically assess blood pressure or physical activity (e.g., contact sports), or any marker of blood-brain-barrier permeability and inflammation, although their associations with NfL levels have been shown in previous works.^35,48^ Nonetheless, we excluded all participants with traumatic brain injury (including concussion or contusion), loss of consciousness, or a history of known neurological, cardiovascular, or autoimmune disorders. Moreover, alterations in the blood-brain-barrier permeability and in the inflammatory response have been mostly suggested after acute cocaine intake, but evidence of long-lasting changes of these markers in CU are still lacking.^49,50^ Importantly, NfL levels in our sample were associated with cocaine use in the last four months but not with use in the last here days (assessed by urine toxicology), which should be the case if NfL elevation was mostly driven by acute or subacute effects.

Taken together, our findings suggest a dose–response relationship of cocaine-related neurotoxicity on neuroaxonal structures. The sensitivity, reliability and low-invasivity of NfL make it an ideal candidate for longitudinal monitoring of active brain pathology in patients with chronic cocaine use. Accordingly, we also suggest considering illicit substance use, specifically of stimulants such as cocaine, as a significant confounding variable in investigations of NfL levels in all neurological and psychiatric studies and in its diagnostic application.

## Data Availability

All data produced in the present study are available upon reasonable request to the authors

## Acknowledgements

We are grateful to Monika Näf, Chantal Kunz, Marlon Nüscheler, Selina Maisch, Jocelyn Waser, Anna Burkert, Meret Speich, Maxine de Ven, Zoé Dolder, Zoe Hillmann, Jessica Grub, and Priska Cavegn for their excellent support with recruitment and assessment of the participants and to Dou Zhiwei and Torsten Hothorn for their much-appreciated advice on statistical methods.

## References

1. Hall MG, Alhassoon OM, Stern MJ, et al. Gray matter abnormalities in cocaine versus methamphetamine-dependent patients: a neuroimaging meta-analysis. The American Journal of Drug and Alcohol Abuse 2015;41:290–299.

2. He Q, Li D, Turel O, Bechara A, Hser Y-I. White matter integrity alternations associated with cocaine dependence and long-term abstinence: Preliminary findings. Behavioural Brain Research 2020;379:112388.

3. Rabin RA, Mackey S, Parvaz MA, et al. Common and gender-specific associations with cocaine use on gray matter volume: Data from the ENIGMA addiction working group. Hum Brain Mapp 2020.

4. Tondo LP, Viola TW, Fries GR, et al. White matter deficits in cocaine use disorder: convergent evidence from in vivo diffusion tensor imaging and ex vivo proteomic analysis. Transl Psychiatry 2021;11:252.

5. Ersche KD, Williams GB, Robbins TW, Bullmore ET. Meta-analysis of structural brain abnormalities associated with stimulant drug dependence and neuroimaging of addiction vulnerability and resilience. Current Opinion in Neurobiology 2013;23:615–624.

6. Parvaz MA, Moeller SJ, D’Oleire Uquillas F, et al. Prefrontal gray matter volume recovery in treatment-seeking cocaine-addicted individuals: a longitudinal study. Addiction Biology 2017;22:1391–1401.

7. Hirsiger S, Hänggi J, Germann J, et al. Longitudinal changes in cocaine intake and cognition are linked to cortical thickness adaptations in cocaine users. NeuroImage: Clinical 2019;21:101652.

8. Jedema HP, Song X, Aizenstein HJ, et al. Long-Term Cocaine Self-administration Produces Structural Brain Changes That Correlate With Altered Cognition. Biological Psychiatry 2021;89:376–385.

9. Makris N, Gasic GP, Kennedy DN, et al. Cortical Thickness Abnormalities in Cocaine Addiction—A Reflection of Both Drug Use and a Pre-existing Disposition to Drug Abuse? Neuron 2008;60:174–188.

10. Vonmoos M, Hirsiger S, Preller KH, et al. Cognitive and neuroanatomical impairments associated with chronic exposure to levamisole-contaminated cocaine. Translational Psychiatry 2018;8.

11. Conrad F, Hirsiger S, Winklhofer S, et al. Use of levamisole-adulterated cocaine is associated with increased load of white matter lesions. J Psychiatry Neurosci 2021;46:E281–e291.

12. Crunelle CL, Kaag AM, Van Wingen G, et al. Reduced frontal brain volume in non-treatment-seeking cocaine-dependent individuals: exploring the role of impulsivity, depression, and smoking. Frontiers in human neuroscience 2014;8:7.

13. Liu Y-L, Bavato F, Chung A-N, et al. Neurofilament light chain as novel blood biomarker of disturbed neuroaxonal integrity in patients with ketamine dependence. The World Journal of Biological Psychiatry 2021:1–9.

14. Bavato F, Cathomas F, Klaus F, et al. Altered neuroaxonal integrity in schizophrenia and major depressive disorder assessed with neurofilament light chain in serum. Journal of Psychiatric Research 2021;140:141–148.

15. Kluwe-Schiavon B, Kexel A, Manenti G, et al. Sensitivity to gains during risky decision-making differentiates chronic cocaine users from stimulant-naïve controls. Behavioural Brain Research 2020;379:112386.

16. Volkow ND, Koob G, Baler R. Biomarkers in Substance Use Disorders. ACS Chemical Neuroscience 2015;6:522–525.

17. Yuan A, Rao MV, Veeranna Nixon RA. Neurofilaments and Neurofilament Proteins in Health and Disease. Cold Spring Harb Perspect Biol 2017;9.

18. Leppert D, Kuhle J. Blood neurofilament light chain at the doorstep of clinical application. Neurol Neuroimmunol Neuroinflamm 2019;6:e599.

19. Khalil M, Teunissen CE, Otto M, et al. Neurofilaments as biomarkers in neurological disorders. Nature Reviews Neurology 2018;14:577–589.

20. Kuhle J, Kropshofer H, Haering DA, et al. Blood neurofilament light chain as a biomarker of MS disease activity and treatment response. Neurology 2019;92:e1007–e1015.

21. Vonmoos M, Eisenegger C, Bosch OG, et al. Improvement of emotional empathy and cluster B personality disorder symptoms associated with decreased cocaine use severity. Frontiers in psychiatry 2019;10:213.

22. Vonmoos M, Hulka LM, Preller KH, Minder F, Baumgartner MR, Quednow BB. Cognitive Impairment in Cocaine Users is Drug-Induced but Partially Reversible: Evidence from a Longitudinal Study. Neuropsychopharmacology 2014;39:2200–2210.

23. Liu HM, Hsieh WJ, Yang CC, Wu VC, Wu KD. Leukoencephalopathy induced by levamisole alone for the treatment of recurrent aphthous ulcers. Neurology 2006;67:1065–1067.

24. Vitt JR, Brown EG, Chow DS, Josephson SA. Confirmed case of levamisole-associated multifocal inflammatory leukoencephalopathy in a cocaine user. J Neuroimmunol 2017;305:128–130.

25. 25. Kexel A-K, Kluwe-Schiavon B, Baumgartner MR, et al. Cue-induced cocaine craving enhances psychosocial stress and vice versa in chronic cocaine users. medRxiv 2022:2022.2001.2013.22268894.

26. Cooper GAA, Kronstrand R, Kintz P. Society of Hair Testing guidelines for drug testing in hair. Forensic Science International 2012;218:20–24.

27. Quednow BB, Steinhoff A, Bechtiger L, Ribeaud D, Eisner M, Shanahan L. High Prevalence and Early Onsets: Legal and Illegal Substance Use in an Urban Cohort of Young Adults in Switzerland. European Addiction Research 2021:1–13.

28. APA E. Diagnostic and statistical manual of mental disorders, Text Revision (DSM-IV-TR). Washington, DC 2000.

29. Beck AT. An Inventory for Measuring Depression. Archives of General Psychiatry 1961;4:561.

30. Bernstein DP, Stein JA, Newcomb MD, et al. Development and validation of a brief screening version of the Childhood Trauma Questionnaire. Child Abuse & Neglect 2003;27:169–190.

31. Rösler M, Retz W, Retz-Junginger P, et al. [Tools for the diagnosis of attention-deficit/hyperactivity disorder in adults. Self-rating behaviour questionnaire and diagnostic checklist]. Nervenarzt 2004;75:888–895.

32. Quednow BB, Kühn K-U, Hoenig K, Maier W, Wagner M. Prepulse Inhibition and Habituation of Acoustic Startle Response in Male MDMA (‘Ecstasy’) Users, Cannabis Users, and Healthy Controls. Neuropsychopharmacology 2004;29:982–990.

33. Scholz C, Cabalzar J, Kraemer T, Baumgartner MR. A Comprehensive Multi-Analyte Method for Hair Analysis: Substance-Specific Quantification Ranges and Tool for Task-Oriented Data Evaluation. Journal of Analytical Toxicology 2020.

34. Hoelzle C, Scheufler F, Uhl M, Sachs H, Thieme D. Application of discriminant analysis to differentiate between incorporation of cocaine and its congeners into hair and contamination. Forensic Science International 2008;176:13–18.

35. Barro C, Chitnis T, Weiner HL. Blood neurofilament light: a critical review of its application to neurologic disease. Ann Clin Transl Neurol 2020;7:2508–2523.

36. Jones PR. A note on detecting statistical outliers in psychophysical data. Attention, Perception, & Psychophysics 2019;81:1189–1196.

37. Vonmoos M, Hulka LM, Preller KH, et al. Cognitive dysfunctions in recreational and dependent cocaine users: role of attention-deficit hyperactivity disorder, craving and early age at onset. British Journal of Psychiatry 2013;203:35–43.

38. Scholz C, Quednow BB, Herdener M, Kraemer T, Baumgartner MR. Cocaine Hydroxy Metabolites in Hair: Indicators for Cocaine Use Versus External Contamination⍰. Journal of Analytical Toxicology 2019;43:543–552.

39. Cantó E, Barro C, Zhao C, et al. Association Between Serum Neurofilament Light Chain Levels and Long-term Disease Course Among Patients With Multiple Sclerosis Followed up for 12 Years. JAMA Neurology 2019;76:1359.

40. Pereira RB, Andrade PB, Valentão P. A Comprehensive View of the Neurotoxicity Mechanisms of Cocaine and Ethanol. Neurotoxicity Research 2015;28:253–267.

41. Guha P, Harraz MM, Snyder SH. Cocaine elicits autophagic cytotoxicity via a nitric oxide-GAPDH signaling cascade. Proceedings of the National Academy of Sciences 2016;113:1417–1422.

42. Beitner-Johnson D, Guitart X, Nestler E. Neurofilament proteins and the mesolimbic dopamine system: common regulation by chronic morphine and chronic cocaine in the rat ventral tegmental area. The Journal of Neuroscience 1992;12:2165–2176.

43. Kovacs K, Lajtha A, Sershen H. Effect of nicotine and cocaine on neurofilaments and receptors in whole brain tissue and synaptoneurosome preparations. Brain Res Bull 2010;82:109–117.

44. Little KY, Ramssen E, Welchko R, Volberg V, Roland CJ, Cassin B. Decreased brain dopamine cell numbers in human cocaine users. Psychiatry Research 2009;168:173–180.

45. Beste C, Stock AK, Zink N, Ocklenburg S, Akgün K, Ziemssen T. How minimal variations in neuronal cytoskeletal integrity modulate cognitive control. Neuroimage 2019;185:129–139.

46. Treadwell SD, Robinson TG. Cocaine use and stroke. Postgraduate Medical Journal 2007;83:389–394.

47. Gattringer T, Pinter D, Enzinger C, et al. Serum neurofilament light is sensitive to active cerebral small vessel disease. Neurology 2017;89:2108–2114.

48. Uher T, McComb M, Galkin S, et al. Neurofilament levels are associated with blood-brain barrier integrity, lymphocyte extravasation, and risk factors following the first demyelinating event in multiple sclerosis. Mult Scler 2021;27:220–231.

49. Barr JL, Brailoiu GC, Abood ME, Rawls SM, Unterwald EM, Brailoiu E. Acute cocaine administration alters permeability of blood-brain barrier in freely-moving rats— Evidence using miniaturized fluorescence microscopy. Drug and Alcohol Dependence 2020;206:107637.

50. Ersche KD, Döffinger R. Inflammation and infection in human cocaine addiction. Current Opinion in Behavioral Sciences 2017;13:203–209.

